# Quantitative differentiation of minimal-fat angiomyolipomas from renal cell carcinomas using grating-based x-ray phase-contrast computed tomography: an ex vivo study

**DOI:** 10.1101/2022.12.06.22283181

**Authors:** Lorenz Birnbacher, Margarita Braunagel, Marian Willner, Mathias Marschner, Fabio De Marco, Manuel Viermetz, Sigrid Auweter, Susan Notohamiprodjo, Katharina Hellbach, Mike Notohamiprodjo, Michael Staehler, Daniela Pfeiffer, Maximilian F. Reiser, Franz Pfeiffer, Julia Herzen

**Affiliations:** Chair of Biomedical Physics, School of Natural Sciences & Department of Physics, Munich Institute of Biomedical Engineering, Technical University of Munich, Germany; Institute of Diagnostic and Interventional Radiology, School of Medicine & Klinikum rechts der Isar, Technical University of Munich, Germany; Institute of Clinical Radiology, Ludwig-Maximilians-University Hospital Munich, Germany; Department of Physics, University of Trieste, Italy; Department of Nuclear Medicine, School of Medicine & Klinikum rechts der Isar, Technical University of Munich, Germany; Department of Diagnostic and Interventional Radiology, University Hospital of Heidelberg, Ruprecht-Karls-University Heidelberg, Germany; Institute of Urology, Ludwig-Maximilians-University Hospital Munich, Germany; Institute of Advanced Study, Technical University of Munich, Germany

## Abstract

**Background:** The differentiation of minimal-fat—or low-fat—angiomyolipomas from other renal lesions is clinically challenging in conventional computed tomography. In this work, we have assessed the potential of grating-based x-ray phase-contrast computed tomography (GBPC-CT) for visualization and quantitative differentiation of minimal-fat angiomyolipomas (mfAMLs) and oncocytomas from renal cell carcinomas (RCCs) on ex vivo renal samples.

**Materials and methods:** Laboratory GBPC-CT was performed at 40 kVp on 28 ex vivo kidney specimens including five angiomyolipomas with three minimal-fat (mfAMLs) and two high-fat (hfAMLs) subtypes as well as three oncocytomas and 20 RCCs with eight clear cell (ccRCCs), seven papillary (pRCCs) and five chromophobe RCC (chrRCC) subtypes. Quantitative values of conventional Hounsfield units (HU) and phase-contrast Hounsfield units (HUp) were determined and histogram analysis was performed on GBPC-CT and grating-based attenuation-contrast computed tomography (GBAC-CT) slices for each specimen. For comparison, the same specimens were imaged at a 3T magnetic resonance imaging (MRI) scanner.

**Results:** We have successfully matched GBPC-CT images with clinical MRI and histology, as GBPC-CT presented with increased soft tissue contrast compared to absorption-based images. GBPC-CT images revealed a significant (p<0.05) difference between mfAML samples (58±4 HUp), hfAML (−58±17 HUp) and RCCs (ccRCCs: 40±12 HUp; pRCCs: 43±9 HUp; chrRCCs: 40±7 HUp) in contrast to corresponding laboratory attenuation-contrast CT and clinical MRI. Due to the heterogeneity and lower signal of oncocytomas (44±10 HUp), quantitative differentiation of the samples based on HUp or in combination with HUs was not possible.

**Conclusions:** GBPC-CT allows quantitative differentiation of minimal-fat angiomyolipomas from oncocytomas and RCCs in contrast to absorption-based imaging and clinical MRI.

## Introduction

In diagnostic imaging, the number of identified renal lesions has been increasing in the past decades as a result of rising incidental detection of particular small renal masses with improved imaging techniques like computed tomography (CT) [1]. Renal lesions are a heterogeneous group of tumors ranging from benign lesions to malignant RCCs. The three most common RCC subtypes are clear cell carcinoma, papillary and chromophobe RCCs presenting with different imaging features, management and prognosis [2, 3]. Often, RCCs display with intratumoral necrosis or cystic lesions [4]. In surgical series after partial or total nephrectomy of radiologically suspect renal lesions, about 80% were malignant RCCs and about 14–20% were benign tumors [5].

The most common benign lesions are renal AMLs and oncocytomas. AMLs consist of variable amounts of adipose tissue, smooth muscle cells and blood vessels [4]. In general, there are two distinct types of AMLs: high-fat and minimal-fat AMLs. A typical hfAML can be diagnosed with great accuracy on ultrasound, unenhanced computed tomography or chemical-shift MRI by detecting macroscopic fat components [6]. However, mfAMLs account for 5% of all renal AMLs and differ from typical hfAMLs due to the lack of detectable macroscopic fat on unenhanced CT, chemical-shift MR images or T2-weighted MR images [7]. On CT or MRI, mfAMLs can mimic RCCs—especially the papillary subtype—leading to unnecessary surgery. Previous studies have analyzed different strategies for discerning mfAMLs from RCCs through imaging with unenhanced [8] and multiphasic CT [9], CT histogram analysis [10, 11], texture analysis [12, 13] as well as chemical-shift [14] and T2 signal ratio on MR images [14, 15]. Increased absolute and relative density of mfAMLs compared to RCCs on unenhanced CT were presented in several works [6, 9, 16, 17], but hemorrhagic or proteinous cysts as well as lymphomas can also display as hyperdense signals on unenhanced CT scans. This overlap in density values of mfAMLs and other renal masses still limits considerably the diagnostic utility of CT [8].

Renal oncocytoma represents about 7 % of all surgically removed kidney masses [4]. Histologically, oncocytomas usually consist of oncocytes, stroma and capillaries, but can also contain central scars and intratumoral hemorrhage. Oncocytomas are often described as a well-demarcated, homogeneously enhancing tissue in contrast-enhanced CTs with a characteristic central scar without calcifications or necrosis. But in several studies, only a small proportion of these tumors showed these characteristic imaging features [4]. Therefore, qualitative and quantitative analysis displays a considerable overlap with ccRCCs in different studies [13, 18, 19]. Preoperative differentiation of benign mfAMLs and oncocytomas from malignant RCCs has a great importance for preventing overtreatment [20], although biopsy or resection due to inconclusive non-invasive preoperative imaging remains the standard of reference.

Grating-based phase-contrast computed tomography (GBPC-CT) is a three-dimensional imaging method visualizing the phase shift induced by an x-ray beam when passing through tissue. This phase shift enables visualization of subtle soft tissue differences in contrast to corresponding conventional attenuation-based CT. While there exist several techniques to access the x-ray phase-contrast signal [21], GBPC-CT is one phase-contrast imaging technique which has been successfully translated to incoherent polychromatic x-ray sources such as clinical high-flux x-ray sources [22]. Previous studies have shown an increased soft tissue contrast of grating-based phase-contrast compared to attenuation-based images [23], e.g. in breast cancer samples [24–26], characterization of atherosclerotic plaque [27, 28], in different renal cysts [29] and RCCs [30]. Moreover, GBPC-CT enables quantitative imaging since the phase-contrast signal can be related to the electron density [31].

In this ex vivo laboratory study, we have evaluated GBPC-CT imaging for a qualitative and most importantly quantitative differentiation of mfAMLs and oncocytomas from RCCs.

## Materials and methods

### Sample acquisition and preparation

The local ethics committee (Ethikkomission der Universität München, Munich) approved this retrospective experimental ex vivo study conducted in accordance with the International Declaration of Helsinki. Written informed constent was acquired by all 28 patients. The total number of human renal ex vivo samples was 28 and comprised eight clear cell (ccRCC; mean size: 9 cm), seven papillary (pRCC; mean size: 12 cm) and five chromophobe RCCs (chrRCC; mean size: 7 cm), three oncocytomas (mean size: 5 cm) after total nephrectomy and five angiomyolipomas (three mfAMLs and two hfAMLs; mean size: 7 cm). Partial or total nephrectomy was performed following the recommendation of the interdisciplinary tumor board. Experienced pathologists selected representative tumor tissue samples of 3 cm maximum diameter and 10 cm maximum length, which were placed in 50 ml plastic containers filled with 4% formaldehyde solution for fixation. In a prior study [30], we reported on the qualitative characterization of 20 RCC samples which are included in the current study.

### Clinical CT and MR imaging

After renal sample collection, preoperative clinical computed tomography data of the corresponding patients were searched in the clinical database. 13 out of 28 patients with a renal tumor had a preoperative CT at 100–120 kVp (unenhanced, arterial, venous and delayed phase), two patients underwent an MRI, one patient was imaged using ultrasound and 13 patients had no imaging in the database. The mean age of patients with RCC was 62 years (range 36–89 years), with oncocytoma 61 years (range 53–77 years) and with angiomyolipoma 36 years (range 9–70 years). Representative measurements of Hounsfield units (HU) in selected tumor masses on unenhanced CT scans were done with placement of regions of interest (ROIs) labeling the entire tumor using the open access software OsiriX (OsiriX 5.8, Pixmeo SARL, Geneva, Switzerland).

All 28 excised samples were also imaged with a 3T-MRI system (MAGNETOM Skyra, Siemens Healthineers, Erlangen, Germany) after placement in a 16-channel hand wrist coil in transversal sequences without the use of contrast media. Detailed MRI sequence parameters can be found in Braunagel et al. [30].

### Grating-based phase-contrast CT

Three complementary image signals can be accessed with a laboratory grating-based phase-contrast computed tomography setup, namely the conventional attenuation image, the phase-contrast signal and the dark-field image [22]. While the phase-contrast signal visualizes subtle soft tissue differences based on changes in electron density, the dark-field signal shows small-angle scattering originating from structures below the physical pixel size [32]. All three contrast signals are simultaneously acquired, can be used for tomographic imaging and are intrinsically perfectly registered. We refer to previous work on details on the used preclinical GBPC-CT setup operating at 40 kVp [26, 30, 33]. While the grating-based attenuation-contrast computed tomography (GBAC-CT) data can be converted to conventional Hounsfield units (HU), so-called phase-contrast Hounsfield units (HUp) can be calculated with the grating-based phase-contrast computed tomography (GBPC-CT) signal [34, 35].

### Histology

After imaging, all samples were sliced into 5-mm thick larger pieces, embedded in paraffin, and further cut with a microtome into 5-μm-thick representative tissue sections. Standard protocols for hematoxylin and eosin (HE) staining based on 10–12 histological slices for each sample were used [30]. Histological workup and diagnoses of all renal tumors based on microscopic evaluation according to histopathological diagnostic guidelines was performed by experienced pathologists [36]. Further staining was applied for the diagnosis of chrRCC using Hale’s staining. The HMB-45 monoclonal antibody allowed for immunohistochemistry staining of hf- and mfAMLs [37].

### Data analysis

An open access DICOM viewing software (OsiriX 5.8, Pixmeo SARL, Geneva, Switzerland) was used by experienced radiologists for image analysis. The histopathologic diagnosis was withheld from the radiologists prior to assessment [30]. In a consensus of radiologists and pathologists, imaging results and histological slices were compared based on features like fat, calcifications or tumor delineation [30]. Ten regions of interest (ROIs) on different transversal slices were placed in each sample (n=28) labeling the entire tumor while excluding surrounding tissues. Mean values and standard deviation of quantitative phase-contrast Hounsfield units (HUp) and conventional attenuation HU values of tumor areas were calculated based on all voxels included in the ROIs for all samples.

Determination of mean values and standard deviations, histogram analysis of HUp and HU values as well as independent t-tests were executed with Excel 2016 (Microsoft, Redmond (WA), USA). For further statistical analysis, IBM SPSS 23.0 (IBM, Armonk (NY), USA) was applied. In this study, we assumed that a p-value of less than 0.05 is of statistically significance [30].

## Results and discussion

Representative slices of clinical CT scans for each tumor subtype with exemplary HU measurements in tumor regions are shown in Fig. 1. Qualitative and quantitative analysis of the images (Fig. 1 B) reveals that while hfAML can be confidently identified (p < 0.05) and distinguished from the other lesion types, it is not possible to correctly differentiate the other tumor subtypes on the clinical CT images. Figure 2 shows representative slices of GBPC-CT, GBAC-CT, T2 fat-saturated MRI as well as histology slices of one mfAML, hfAML and oncocytoma sample, respectively. In contrast to the attenuation signal (GBAC-CT), phase-contrast CT (GBPC-CT) presents with an increased soft tissue contrast with good correlation to histological slices revealing many tissue substructures like smooth muscle cells and blood vessels in AMLs and oncocytes and capillaries in oncocytomas, respectively. T2 MR images also display superior soft tissue contrast compared to GBAC-CT at a significantly lower spatial resolution. The shown mfAML sample appears very homogeneous, which was similarly observed in all mfAML samples (n=3) with high signals in GBPC-CT and homogeneous low signals in T2 MRI. In contrast to the heterogeneous signals of oncocytomas with low and high signal areas in all imaging modalities. Due to macroscopic fat, hfAML appears homogeneous at low signal values (negative HU and HUp-values) on all imaging modalities. Figure 3 shows representative GBPC-CT, GBAC-CT, T2 fat-saturated MRI slices and histology sections of the three major malignant RCC types (ccRCC, pRCC and chrRCC). The RCC samples appear generally with lower signal intensity than the benign samples in Fig. 2 aside from adipose tissue parts. The ccRCC and pRCC samples present heterogeneously with a lower signal than surrounding normal kidney tissue in GBPC-CT, GBAC-CT and T2 MRI. In contrast to ccRCC and pRCC, the chrRCC sample shows a more homogeneous tumor tissue structure with a lower signal compared to surrounding normal kidney in GBPC-CT and higher signal in T2 MRI. Due to low soft tissue contrast of GBAC-CT, the tumor and normal tissue of the chrRCC sample could not be discriminated from surrounding formalin. From Fig. 2 and 3, a clear visual difference between mfAML, hfAML and the other types of tumor can be appreciated on the GBPC-CT images, while the differentiation based on other imaging modalities remains very challenging. This result is highlighted in Fig. 4 on the example of the signal distribution histograms of the phase-contrast signal for the samples presented in Fig. 2 and 3. mfAML is characterized by a high HUp signal with a narrow signal distribution, as compared to other tumor subtypes.

**Fig 1.**
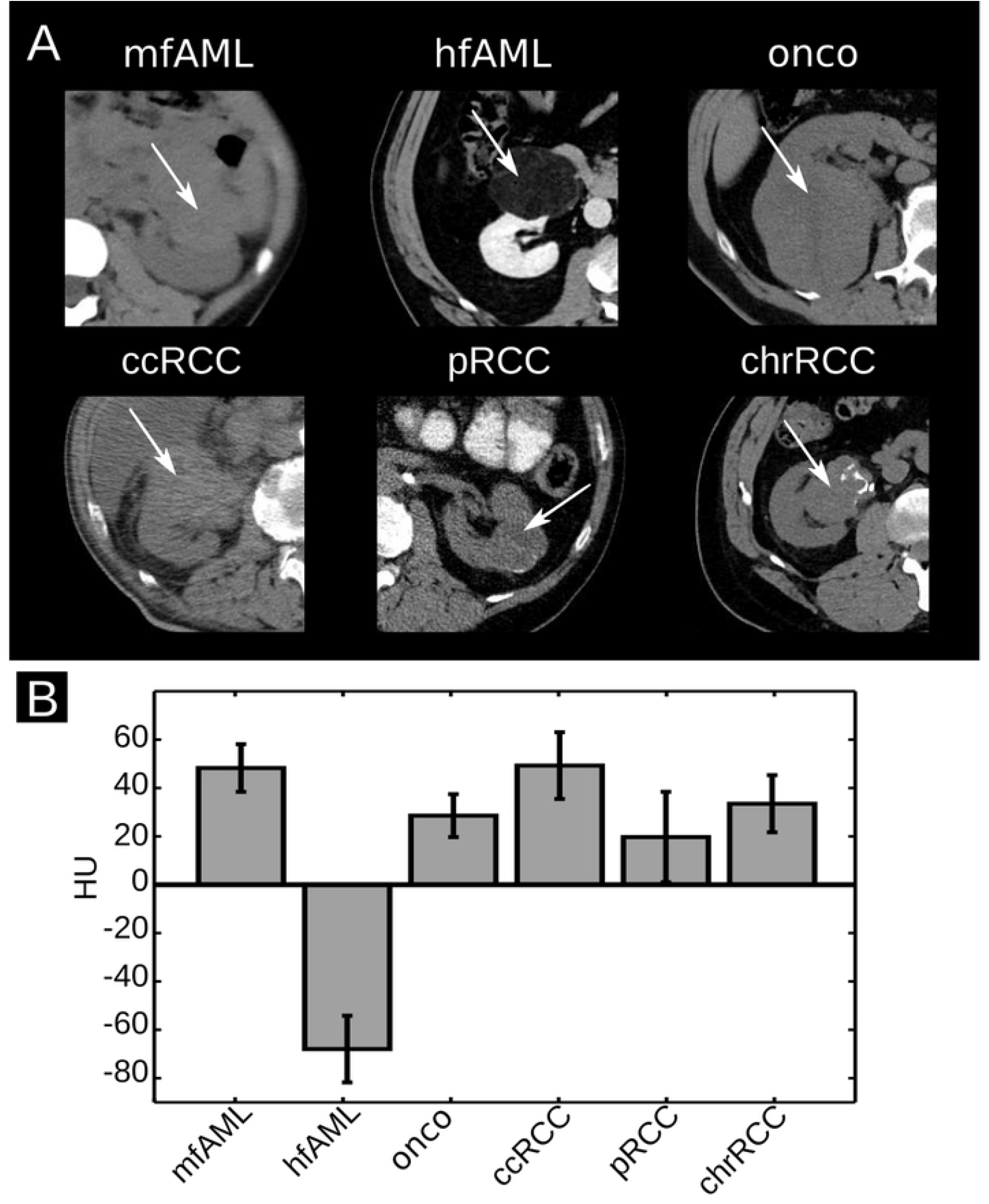
Clinical renal CT slices and distribution of HU values. (A) Representative clinical axial CT slices of kidney lesion samples in soft tissue window. Aside from the hfAML (venous phase CT) all other scans are unenhanced. The arrows highlight the tumorous tissue. (B) Mean and standard deviation of the quantitative HU values from the images presented in (A).

**Fig 2.**
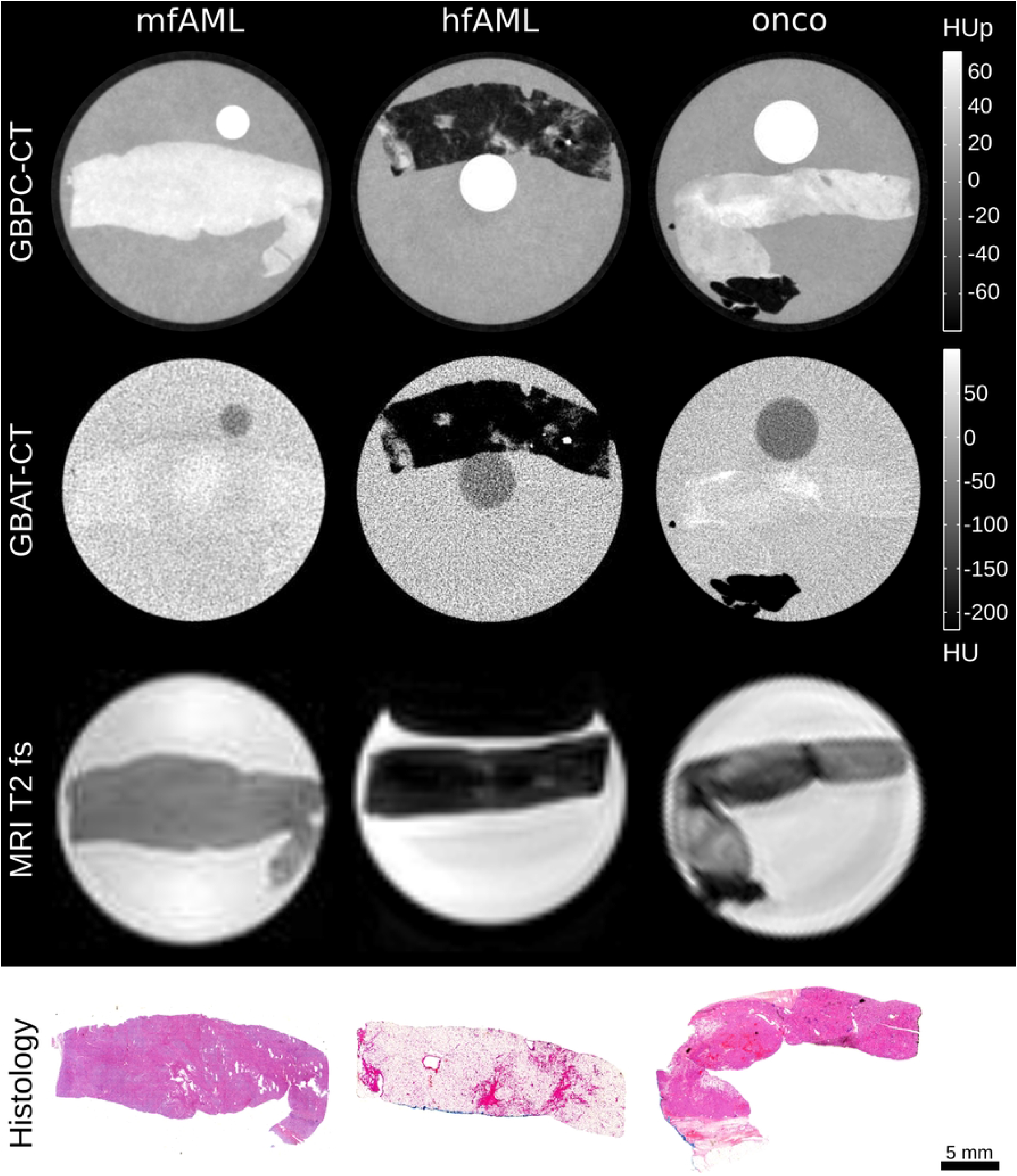
Preclinical imaging results of benign kidney lesions in histopathological correlation. Typical slices of a mfAML, hfAML and oncocytoma (onco) obtained with GBPC-CT, GBAC-CT and T2 fat saturated MRI (MRI T2 fs). The bottom row displays the corresponding histological slices that were obtained for validation with HE-staining.

**Fig 3.**
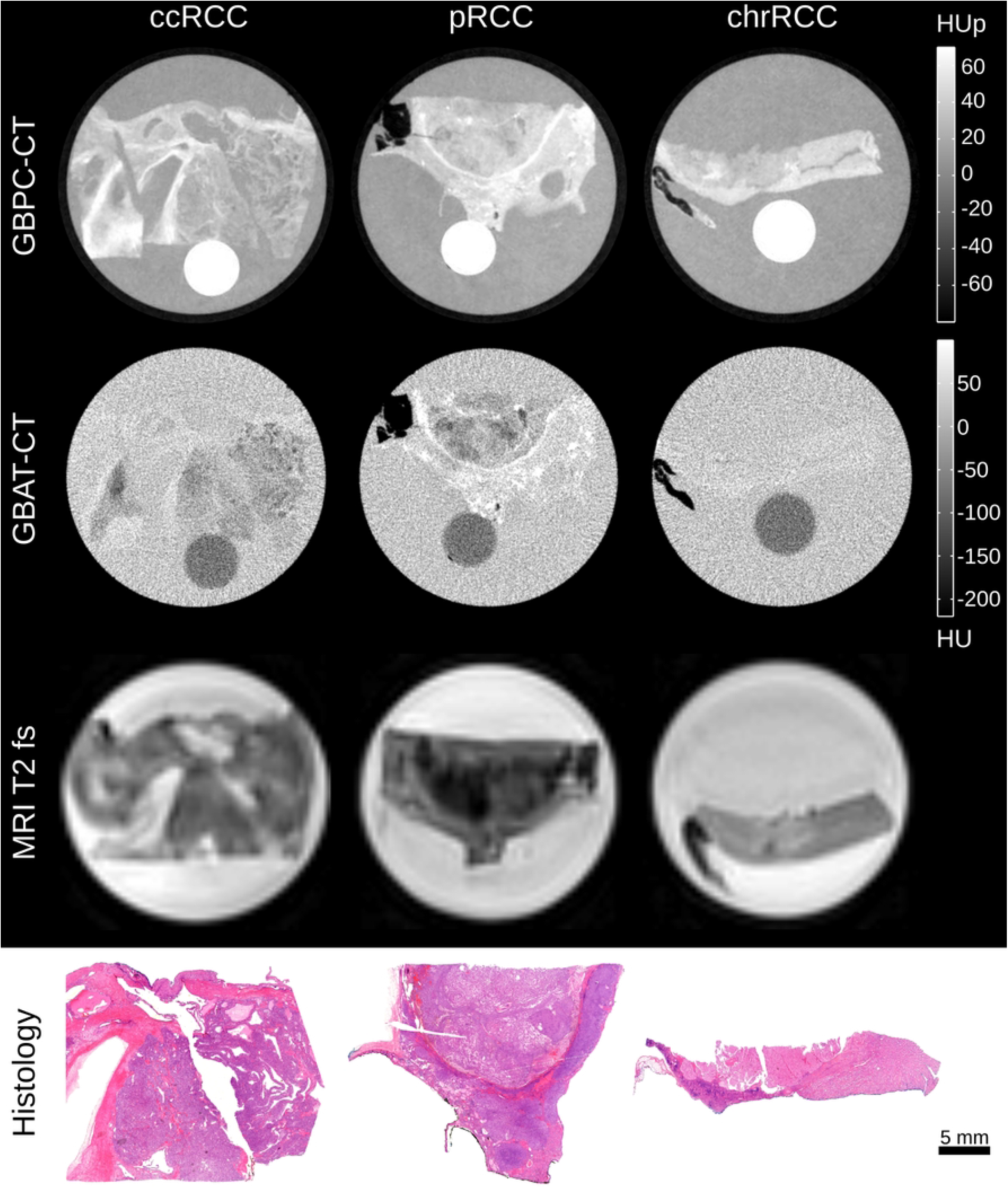
Preclinical imaging results of renal cell carcinomas in histopathological correlation. Representative slices of a ccRCC, pRCC and chrRCC obtained with GBPC-CT, GBAC-CT and T2 fat saturated MRI (MRI T2 fs). The bottom row shows the corresponding histological slices that were obtained for validation with HE-staining.

**Fig 4.**
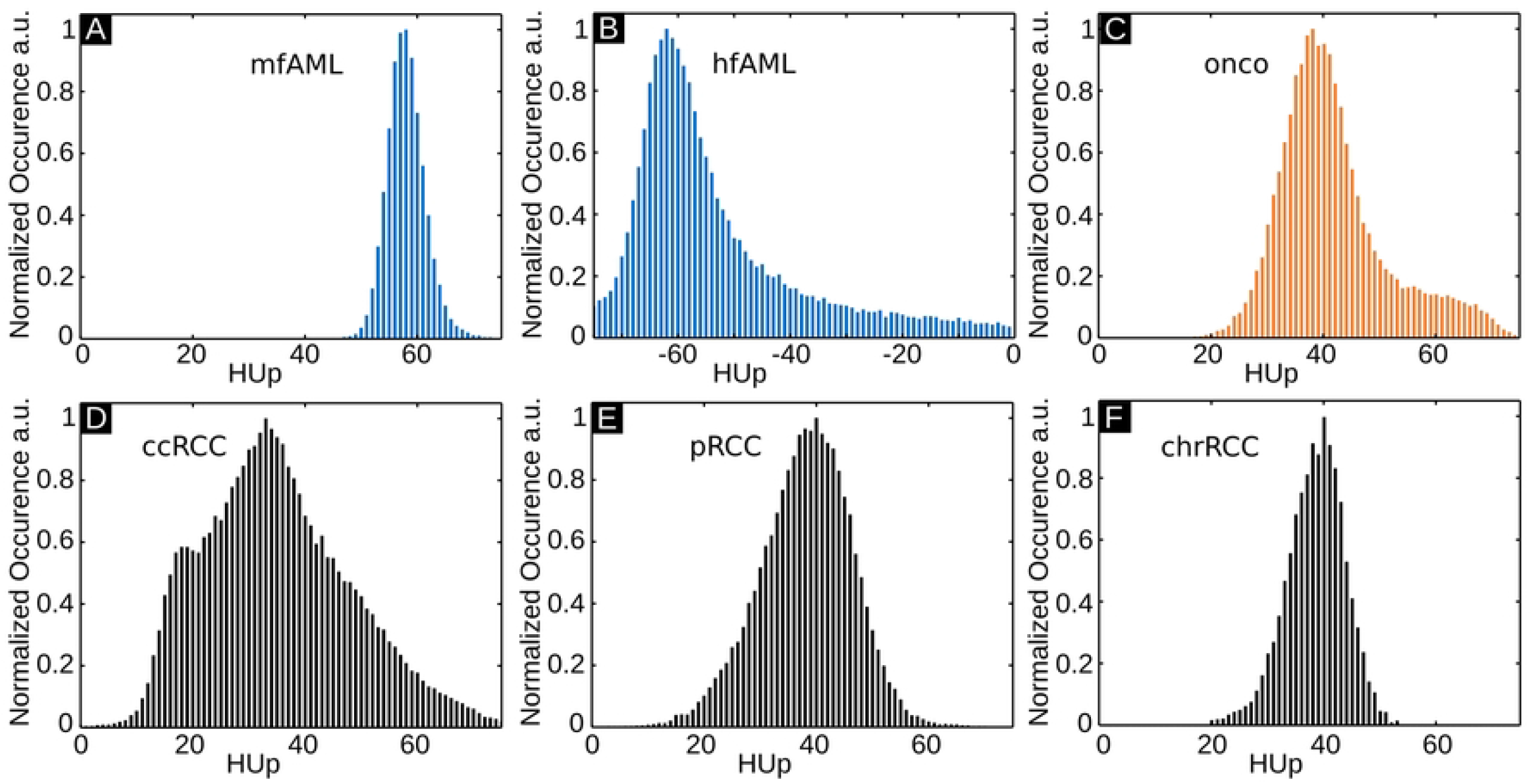
Histogram analysis of quantitative GBPC-CT data. GBPC-CT signal distribution histograms for the samples presented in Fig. 2 and 3. The peak of the mfAML values is quite sharp compared to the other results. The shown range of all histograms is 80 HUp.

Table 1 summarizes the mean signal values with the corresponding standard deviations for all tumor subtypes, as evaluated by placing 10 ROIs on all the samples. Due to the intrinsically perfect registration of GBAC-CT and GBPC-CT images, identical ROIs for both modalities were used. We can observe that it is not possible to discriminate different tumor types based on the attenuation (GBAC-CT) imaging modality alone, which originates in parts from the low signal-to-noise ratio in the GBAC-CT images. This finding correlates well with the results reported in Figure 1 (B). This result is further visualized in Fig. 5 (A) and (B), where boxplots for all the samples are presented. Here, the mean phase-contrast signals of all mfAML samples differ significantly from other tumor subtypes (p<0.05). Moreover, this sample type is characterized by the smallest standard deviation.

**Table 1.**
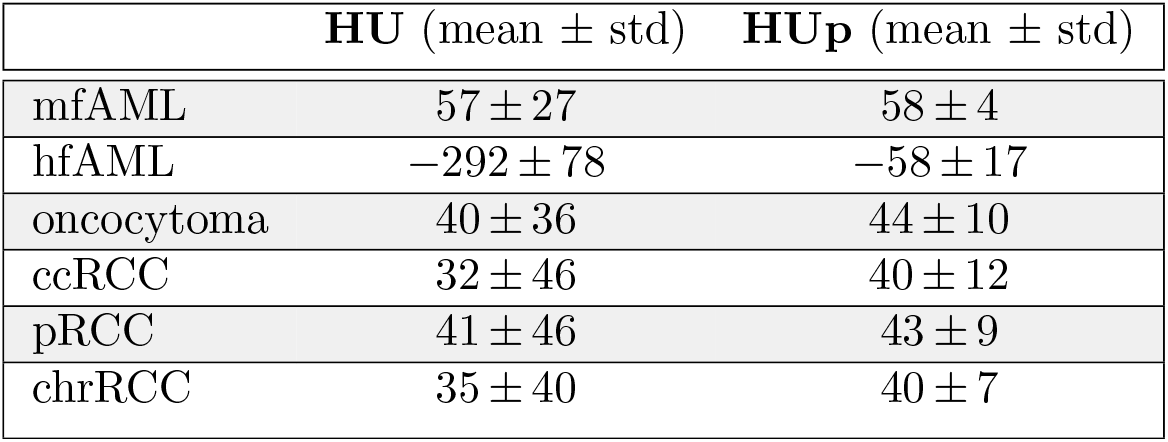
Mean and standard deviation (std) of HU and HUp signals for conventional GBAC-CT (HU) and GBPC-CT (HUp) slices for different types of renal lesions.

**Fig 5.**
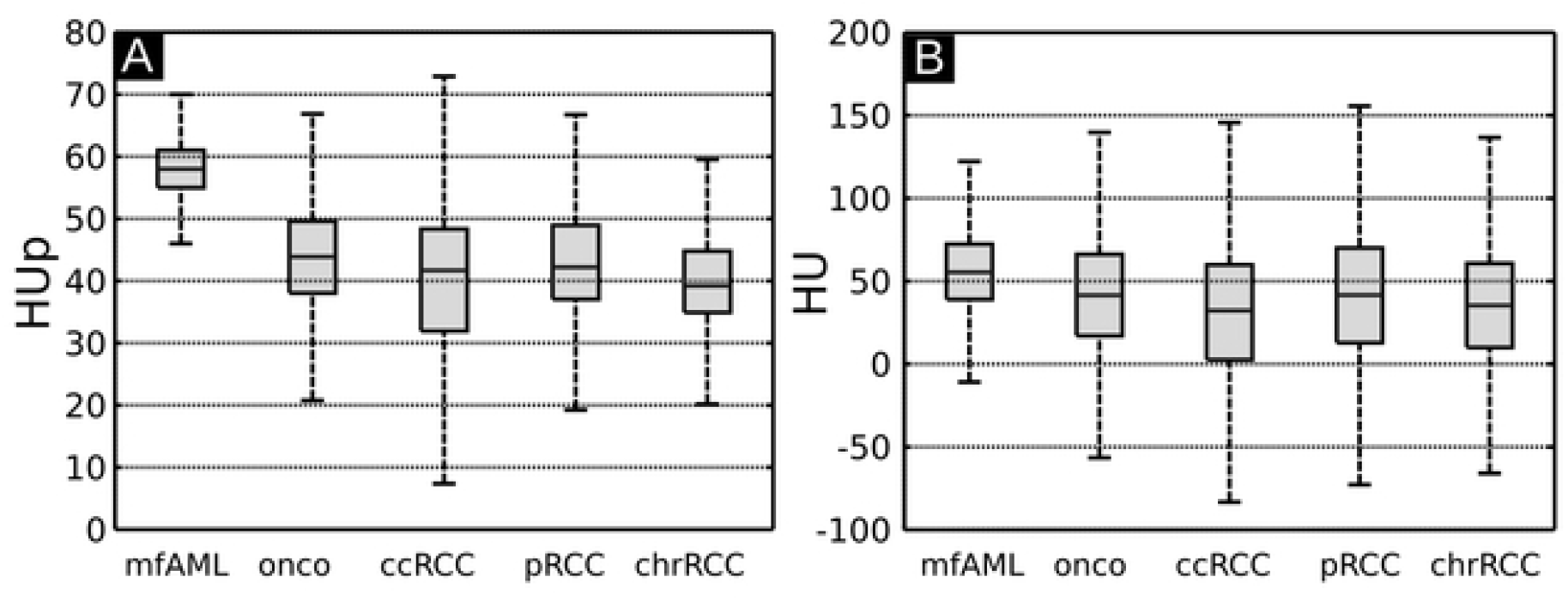
Boxplots of HUp and HU results. Boxplots including all samples of the different tumor subtypes mfAML, oncocytomas (onco), ccRCC, pRCC and chrRCC (n=26). Mean and standard deviation values for GBPC-CT in HUp (A) and for GBAC-CT in HU (B) are shown here. hfAML samples were excluded for visualization reasons.

Contrary to mfAML, hfAMLs exhibit a broader signal distribution due to the presence of different tissue components due to different amounts of fat, smooth muscle cells and vessels in the samples. Quantitatively, our hfAML results are characterized by very low HU and HUp values (− 292±78 HU; − 58±17 HUp) as well as low signal intensities in the MR images, thus hfAML samples can be clearly differentiated from other tumor subtypes in GBAC-CT, GBPC-CT and MR images (p<0.05). Calculation of skewness and curtosis of the signal distribution did not reveal any further significant difference between different types of tumors and is therefore not shown here.

## Conclusion

The results of this laboratory ex vivo study on renal tumor samples indicate that GBPC-CT imaging is better able to differentiate mfAMLs from RCCs and oncocytomas in contrast to GBAC-CT and clinical MRI without contrast media. GBPC-CT revealed significantly higher phase-contrast signals within more homogeneous tissues in mfAMLs compared to RCCs and oncocytomas. In general, the results are in good agreement with the results of a previous study [30], which demonstrated that GBPC-CT yields a superior soft tissue contrast and better delineation of malignant RCC from normal kidney tissue at high spatial resolution. Reliable differentiation of mfAML from different RCC subtypes remains challenging with current clinical tools [11, 14, 38]. Previous studies showed significant differences in absolute density values of mfAMLs especially on unenhanced CT scans [6, 8, 17, 39]. The resulting overlap in density values of mfAML and other renal masses limits diagnostic utility and applicability of density thresholds in the diagnosis of mfAML [8, 40].

In our work, representative unenhanced clinical CT scans of RCCs and benign renal tumors also presented similar density values of ccRCC and mfAML. It was previously shown that multiparametric MR imaging can distinguish between mfAML and the most common renal tumor—the ccRCC—but fail to differentiate between mfAML and pRCC due to same low signal intensities on T2-weighted MR images [15]. Therefore, other imaging techniques should be considered. In the past, it was reported that combining the complementary information obtained from absorption-based and phase-contrast imaging can be used for a better discrimination of different soft tissue [23]. Our study indicates that mfAMLs have a significantly higher quantitative phase-contrast signal within a more homogeneous tissue structure than the three investigated RCC subtypes or the oncocytoma samples. We assume that the higher phase-contrast signal of mfAML specimens can be attributed to the reported increased amount of blood vessels and smooth muscle cells in mfAMLs, which can be related to higher electron density and thus HUp values [4, 31, 41].

While GBAC-CT was acquired at a much higher resolution than clinical CT, GBAC-CT did not yield a significant difference between the mfAML and other tumor subtypes due to high standard deviations (Fig. 5 and Tab. 1). Since the measured attenuation-based CT numbers in Hounsfield units are affected by different kV values due to the energy dependence of the attenuation coefficient, previous studies calculated density ratios of mfAML similar to normal kidneys [8, 39]. In contrast to that, quantitative values of phase-contrast CT are energy-independent as they are based on electron density. Thus, our quantitative measurements of renal samples at 40 kVp of phase-contrast CT imaging can be transferred as reference to clinical applications in the future with higher photon energies [42]. Previously, it has been reported that even at clinically relevant spatial resolution, soft tissue contrast achievable with x-ray phase-contrast is superior to that of attenuation-based CT [30]. Furthermore, phase-contrast Hounsfield units are not strongly influenced by iodine contrast media [29]. Unfortunately, our study revealed that based on GBPC-CT or even on a combination of GBAC-CT with GBPC-CT it is not possible to differentiate oncocytoma from malignant kidney tumors. This originates mainly from the high heterogeneity observed in oncocytoma as well as in RCCs, which correlates with previous studies of unenhanced clinical CT images [4, 18, 38]. Future studies should strive to perform a more advanced signal analysis for a better characterization of the observed heterogeneity for a possible better differentiation.

The presented study has several limitations. Our conclusions are based only on three samples of mfAML. Typical hfAMLs or mfAMLs that are proven by biopsy do not undergo surgery, which limits the availability of the mfAML samples. Future studies should seek to validate the results on a higher number of mfAML samples to substantiate further quantitative assessment of mfAMLs.

Currently, the application of GBPC-CT is limited to preclinical studies and a translation to clinical phase-contrast imaging has to be assessed critically. There are a number of technical challenges that need to be addressed before this technology can be transferred to clinical settings. The main challenges that need to be addressed are the size of the available gratings and thus the field of view, the stability of the interferometer in clinical conditions, the radiation dose and the acquisition time as well as a successful shift to clinical energies. With respect to physical limitations, the benefit of shifting to higher energies was shown in Willner et al. [42] with monochromatic synchrotron sources. Nonetheless, the use of a clinical x-ray source is expected to reduce the performance of GBPC-CT, which has also to be addressed in future work. Recently, a grating interferometer has been integrated into a clinical CT system [44], yet the focus there lies on the dark-field signal, which differs strongly from requirements of visualizing subtle soft tissue differences as realized in the setup used here. In addition, the success of dual-energy and spectral imaging allowing to perform quantitative CT imaging has reduced the need for GBPC-CT in a clinical setting [45].

Meanwhile, GBPC-CT in a preclinical setting could be beneficial for a more sophisticated histopathological workup. GBPC-CT could provide a better understanding of the 3D morphology and quantitative composition of different tissue types at high spatial resolution. This is information that is currently hardly accessible in this combination with conventional histological analysis [23, 46, 47]. However, further increase of GBPC-CT performance is needed to push GBPC-CT towards virtual histology.

## Data Availability

All relevant data are within the manuscript.

## Acknowledgments

We acknowledge financial support through the European Research Council (ERC, H2020, AdG 695045), the DFG Cluster of Excellence Munich-Centre for Advanced Photonics (MAP, EXC158) and the DFG Gottfried Wilhelm Leibniz program. This work was carried out with the support of the Karlsruhe Nano Micro Facility (KNMF, www.kit.edu/knmf), a Helmholtz Research Infrastructure at Karlsruhe Institute of Technology (KIT). We highly appreciate the contribution of the Institute of Pathology at the Ludwig-Maximilians-University Hospital Munich.

## Data availability

All relevant data are within the manuscript. Original data can be accessed on request.

